# Role of genetic risk on progression to diabetes in children with acute pancreatitis

**DOI:** 10.64898/2026.05.23.26353958

**Authors:** Lu Zhang, Faizan Ahmed, Seth A. Sharp, Han Sun, Swaraj Thaman, Clive H. Wasserfall, Anna L. Gloyn, Maisam Abu-El-Haija

## Abstract

**Background:** Acute pancreatitis (AP) is an established risk factor for diabetes, with approximately 20% of children developing either prediabetes or diabetes within one year of their first episode. Little is known about the diabetes pathophysiology or which individuals are at highest risk. We aimed to evaluate whether genetic risk scores (GRS) for type 1 (T1D) and polygenic risk scores (PRS) type 2 diabetes (T2D) are associated with progression to dysglycemia following AP.

**Methods:** Clinical data were available for 123 children (mean age (IQR), 12 (8-15) years; mean body mass index (BMI), 21.8) with AP who were followed for >1 year. Array genotyping coupled with imputation using the TOPMed reference panel was performed. Genetic ancestry was predicted using a random forest classifier. GRS for T1D and T2D were calculated using either an ancestry-appropriate (T1D-GRS) or a multi-ancestry (T2D-PRS) weighted framework. To evaluate risk compared to the population we used predefined GRS thresholds from UK Biobank.

**Results:** Among the 123 subjects, 24 developed dysglycemia (5 with diabetes and 19 with prediabetes). The majority (75.6%, n=93) of children were of European ancestry. Comparison of the T1D-GRS burden with the UK BioBank showed numerically higher proportions for any given threshold. At the top 5% threshold, 9.7% of our cohort were classified as high-risk compared to 5% in UK Biobank (p<0.05). The elevated T1D-GRS could be primarily attributed to non-HLA variants and was more enriched in those testing positive for ≥1 islet-autoantibody. The T2D-PRS was also elevated in the dysglycemic group but only reached statistical significance in those who were obese.

**Conclusion:** These findings highlight the potential role of both T1D-GRS and T2D-PRS in investigating diabetes susceptibility following AP.

## INTRODUCTION

Acute pancreatitis (AP) is increasingly recognized as a risk factor for subsequent diabetes, with studies showing that even a single AP episode increases its risk (1,2). Emerging genetic evidence indicates that genetic risk scores (GRS) for type 1 diabetes (T1D) are predictive of disease development, and polygenic risk scores (PRS) for type 2 diabetes (T2D) can inform diabetes heterogeneity (3). It has been increasingly recognized that diabetes with pancreatitis is a comorbid condition now often described as “Type 3C” or “pancreatogenic diabetes” but the molecular underpinnings of post-pancreatogenic diabetes are poorly understood. The contribution of established genetic risk factors for T1D and T2D has not been assessed in the development of AP.

In this brief report, we address this gap by evaluating T1D-GRS and T2D-PRS, including partitioned components, to determine their utility in predicting which pediatric patients will develop diabetes following a single AP episode and to inform on the contributions of established risk factors for autoimmune and metabolic forms of diabetes.

## METHODS

### Patients

We enrolled 123 pediatric subjects (age: <21 years) with their first AP episode who presented to the Cincinnati Children’s Hospital Medical Center (AP registry, IRB#2021-4050) meeting the inclusion criteria for AP (5). Based on the body mass index (BMI) percentiles, individuals in the cohort were classified as normal weight (<85%), overweight (85-95%), or obese (≥95%). Patients with prediabetes (defined as HbA1c ≥5.7% and/or fasting glucose ≥100 mg/dL) and diabetes (defined as HbA1c ≥6.5% and/or fasting glucose ≥126 mg/dL or non-fasting glucose >200 mg/dL) were grouped in the dysglycemic group. Dysglycemia classification was based on the most recent HbA1c and/or fasting glucose levels within the first year of follow-up.

### Islet autoantibody measurement

Islet autoantibodies (AAb; GADA, IA-2A, and ZnT8A) were quantified using Enzyme-Linked Immunosorbent Assay (ELISA) (Kronus, Star, ID) as previously described (6). Insulin AAb (IAA) levels were measured using a modified luciferase immunoprecipitation system (LIPS) (2). The same validated positivity thresholds were applied: GADA>5 IU/mL, IA-2A>15 IU/mL, ZnT8A>20 IU/mL, and IAA>8074 relative light units (RLU). These assays were performed in a laboratory maintaining rigorous standardization through the Islet AAb Standardization Program (IASP). The sensitivities (sn) and specificities (sp) were as follows: GADA (sn:76%; sp:100%), IA-2A (sn:66%; sp:99%), ZnT8A (sn:80%; sp:98%), and IAA (sn:40%; sp:98%).

### Genotyping quality control and imputation

DNA was extracted from whole blood and genotyped using either Illumina arrays (Omni2.5Exome-8 v1.5, n=50; or Global Diversity Array-8 v1.0, n=73). Rigorous quality control (QC) was performed using standard thresholds. At the variant level, we excluded single nucleotide polymorphisms (SNPs) with a missing call rate >2% (--geno 0.02) and minor allele frequency (MAF) <0.5% (--maf 0.005). Furthermore, markers that significantly deviated from the Hardy-Weinberg Equilibrium (p<10-6) were removed (--hwe 1e-6). At the individual level, samples exhibiting a missing genotype rate >3% were filtered out (--mind 0.03). Genotypes were imputed using the TOPMed Imputation Server (7). Pre-phasing was performed using Eagle (8), as specified in the server pipeline. Imputation was conducted against the TOPMed mixed-ancestry reference panel (r3/Freeze 8) (9) using the GRCh38/hg38 genome build. After imputation, variants were filtered based on an R2 >0.3 threshold to ensure the quality of the imputed genotypes.

### Ancestry inference and calculation of polygenic risk scores

Genetic ancestry was inferred using Genetic Ancestry Prediction (GAP) (10), an in-house tool that employs a random forest classifier to assign the most probable ancestry to each sample. The model was trained and validated using high-coverage whole-genome sequencing data from 2,504 unrelated individuals in the 1000 Genomes Project, demonstrating superior performance over other machine learning architectures (11).

All GRS/PRS calculations were conducted using the PRS Extension for Diabetes Mellitus (PRSedm) software package (12), which allows the simultaneous generation of GRS/PRS for specific ancestries, as well as hard clustering approaches for partitioned scores (13-14). For T1D, we implemented the t1dgrs2-luckett25 model, an updated "GRS2x" PRS framework consisting of 67 variants with HLA interaction and partitioning (12,13). For T2D, we utilized 1,289 cross-ancestry variants for the multi-ancestry cohort and 1,283 variants optimized for European (EUR) populations (12,14). Using a hard clustering framework, the genetic risk was decomposed into eight non-overlapping physiological clusters, collectively referred to as partitioned PRSs (pPRSs), comprising beta-cell function, adipose tissue, obesity, lipodystrophy, liver metabolism, lipid metabolism, metabolic syndrome, and residual glycemic traits. To maintain data integrity, missing variants were automatically addressed using the software’s integrated imputation and backfilling modules. Importantly, all SNP lists used in these models are open-source and directly accessible within the PRSedm package.

### Statistics

Continuous variables were compared using Welch’s t-test to account for potential unequal variance. When the assumption of normality was violated, the Mann-Whitney U test was employed as a non-parametric alternative, specifically for the analysis of BMI (Neg/Pos) data. To facilitate cross-scale comparisons and visualizations, PRS from different scales were standardized to z-scores. Kernel Density Estimation (KDE) plots were generated to visualize the distribution of these standardized scores. All statistical tests were two-sided, and a p-value of <0.05 was considered statistically significant. The expected proportions were defined based on T1D-GRS cutoffs from the UK Biobank EUR cohort, which are publicly accessible via the PRSedm package. Notably, the reference sample size and EUR ancestry composition were aligned with those reported in previous studies (15), ensuring the robustness of the binomial test comparisons between the observed and expected frequencies.

## RESULTS

### Cohort characteristics and genetic ancestry

Among the 123 subjects, 24 (19.5%) developed dysglycemia (5 with diabetes and 19 with pre-diabetes) within the first year of follow-up after their AP episode. The age at first AP episode ranged widely across the cohort, with most cases occurring during adolescence (Table 1). Age and sex distributions were comparable between groups (Table 1). Estimated genetic ancestry was predominantly EUR (n=93, 75.6%), followed by African (AFR, n=17, 13.8%), Admixed American (AMR, n=12, 9.8%), and South Asian (SAS, n=1, 0.8%) representation (Fig. 1A). When stratified by glycemic status, both groups showed similar ancestry distribution patterns (Fig. 1B).

**Figure 1.**
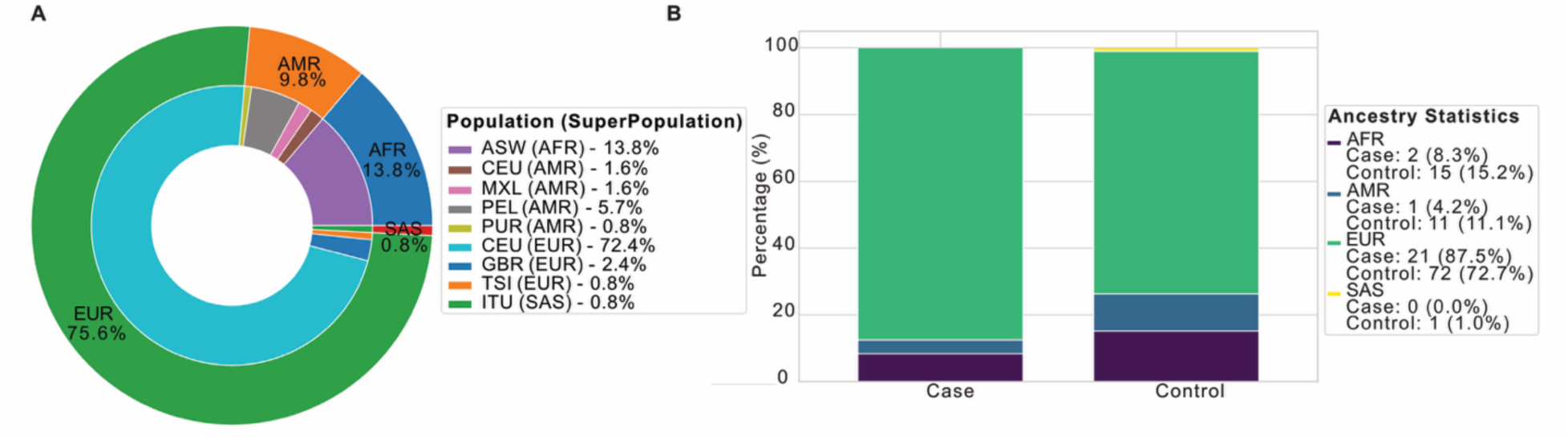
Genetic ancestry distribution. **(A)** Donut chart showing the overall ancestry composition of the cohort, grouped into super populations: European (EUR, 75.6%), African (AFR, 13.8%), Admixed American (AMR, 9.8%), and South Asian (SAS, 0.8%). Inner segments show details of contributing subpopulations. **(B)** Stacked bar plot illustrating the distribution of ancestral backgrounds across dysglycemic cases (prediabetes/diabetes) and normoglycemic controls.

**TABLE 1.** Cohort Demographics.

| Characteristic | Groups |  | p-value |
| --- | --- | --- | --- |
|  | Dysglycemic | Normoglycemic |  |
| **N** | 24 | 99 | - |
| **Age at index AP (years)** | 14.0 (9.0-17.0) | 12.0 (7.0-15.0) | 0.20 |
| **Age at DM (years)** | 14.7 (9.9-17.3) | - | - |
| **Time from AP to DM (days)** | 326.0 | - | - |
| **Sex** |  |  | 0.47 |
| **Male, n (%)** | 14 (58) | 47 (47) |  |
| **Female, n (%)** | 10 (42) | 52 (53) |  |
| **BMI percentile** | 87 (57.5-96.0) | 74 (36.0-94.0) | 0.22 |
| **Overweight, n (%)** | 3 (13) | 16 (16) |  |
| **Obese, n (%)** | 9 (38) | 24 (24) |  |
| **Ethnicity (n)** |  |  | - |
| **Not Hispanic or Latino** | 22 | 94 |  |
| **Hispanic or Latino** | 1 | 4 |  |
| **Unknown** | 1 | 1 |  |
| **Race (n)** |  |  | - |
| **White (n)** | 20 | 86 |  |
| **Black (n)** | 1 | 11 |  |
| **Asian (n)** | 0 | 1 |  |
| **Other/unknown (n)** | 3 | 1 |  |

Data is represented as n, % or median (IQR). Ethnicity and race are self-reported.

### Genetic risk profiles for Type 1 Diabetes

One subject was excluded due to a prior Total Pancreatectomy with Islet Autotransplantation (TPIAT), leaving 122 subjects for analysis. The dysglycemic group showed an elevated global T1D-GRS across all ancestries (Supplementary Fig. 1A), but no significant differences were observed in the HLA or non-HLA partitioned scores (Supplementary Fig. 1B). Furthermore, the T1D-GRS did not significantly correlate with islet AAb status, regardless of the number of positive AAbs (Supplementary Fig. 2). To evaluate whether our cohort carried a higher genetic risk for T1D than the general population, we examined the proportion of individuals exceeding predefined GRS thresholds. We restricted our analysis to individuals of EUR ancestry (n=93) to ensure consistency in the genomic background with reference population. Using cutoffs derived from the UK Biobank as a reference, we observed consistently higher proportions in our cohort across all thresholds. Enrichment at the top 5% threshold reached statistical significance under the binomial test (p<0.05), indicating a significant over-representation of pancreatitis patients among individuals in the highest 5% of the GRS distribution (Fig. 2).

**Figure 2.**
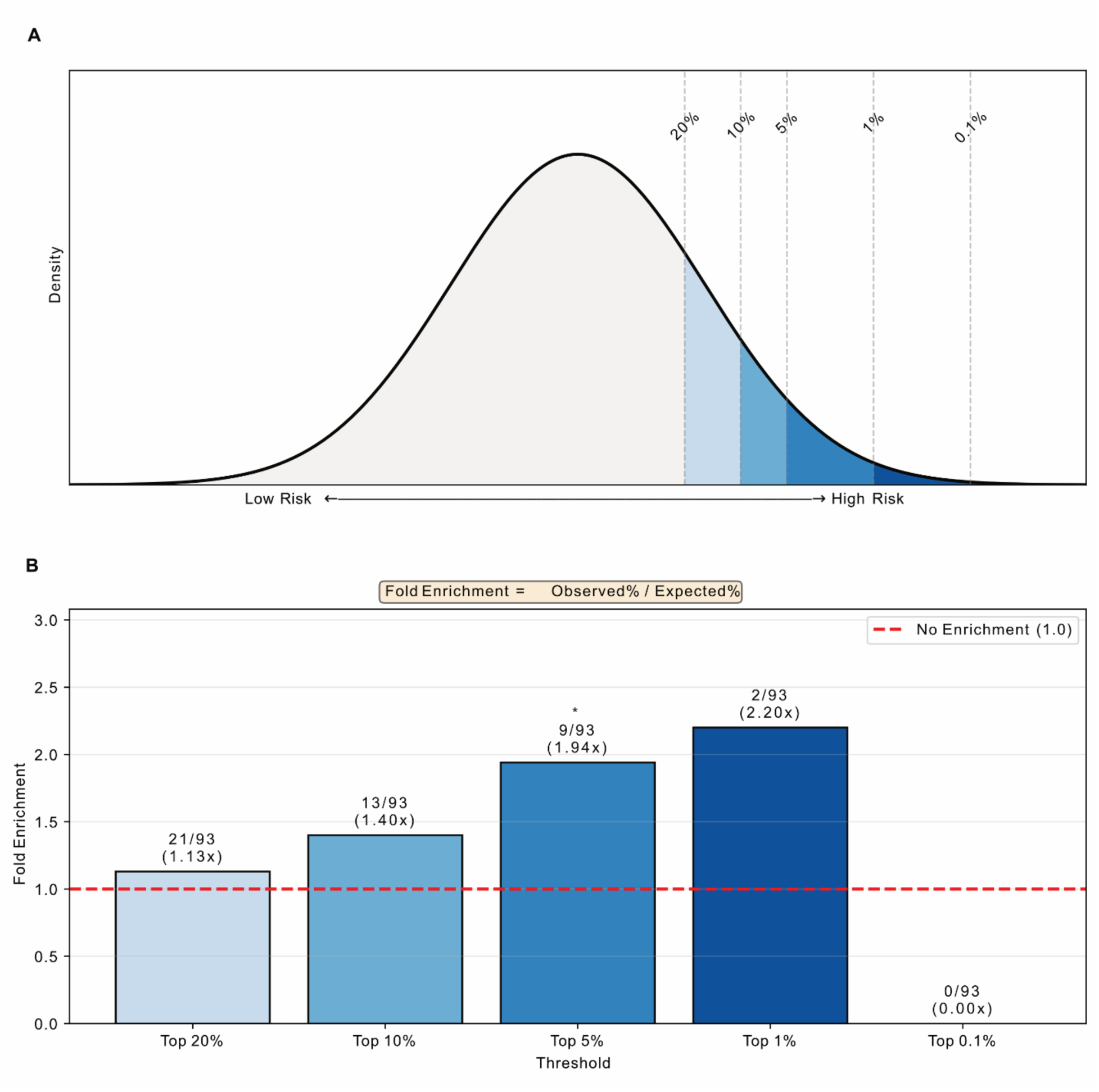
Population Distribution and T1D-GRS Enrichment. **(A)** Theoretical T1D-GRS distribution with threshold region indicated by graduated shading (light to dark: Top 20% to Top 0.1%). **(B)** Case enrichment at each T1D-GRS threshold. Numbers indicate case counts (n/93) and fold enrichment values. Red dashed line represents the baseline (no enrichment). Higher GRS thresholds show increased case enrichment, reaching 2.20-fold at Top 1%.

### Genetic risk profiles for Type 2 Diabetes

Although the global T2D-PRS trended higher in the dysglycemic subjects (Supplementary Fig. 3A), no individual metabolic cluster reached statistical significance (Supplementary Fig. 3B). Whilst the T2D-PRS did not differ by glycemic status in healthy-weight or overweight individuals, it was significantly elevated in dysglycemic subjects within the obese subgroup (p=0.04; Fig. 3). When we limited our analysis to individuals of EUR ancestry, the global T2D-PRS trends remained consistent with our primary findings, and although no individual metabolic clusters reached statistical significance, the residual glycemic cluster exhibited a non-significant elevation in the dysglycemic group (Supplementary Fig. 4).

**Figure 3.**
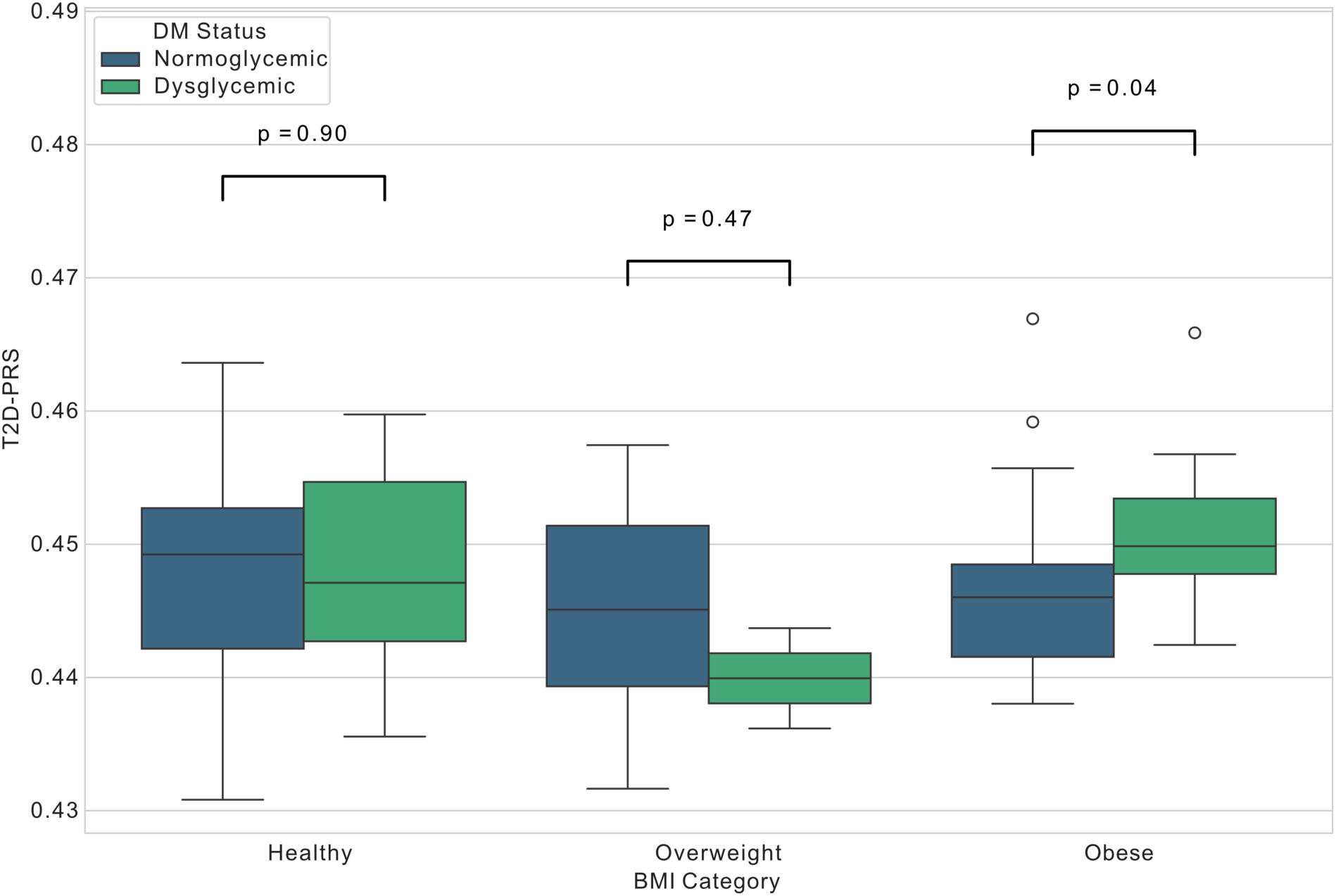
Distribution of T2D-PRS across BMI categories stratified by glycemic status. Data are presented for participants of European ancestry (EUR). Boxplots show T2D-PRS for individuals with normoglycemia (blue) and dysglycemia (green) within healthy weight, overweight, and obese categories. A significant difference in T2D-PRS was observed specifically within the obese group. Sample sizes are as follows: healthy (n=69; 12 dysglycemic, 57 normoglycemic), overweight (n=18; 2 dysglycemic, 16 normoglycemic

## CONCLUSIONS

This study was designed to help us understand the role of established genetic risk factors for diabetes development in a cohort of pediatric subjects with a single episode of AP. Our findings suggest that further studies on the genetic architecture of diabetes in AP, particularly the convergence of diabetes and AP variants, are worth investigating. Our most striking finding was that individuals in our cohort carry a disproportionately higher genetic risk burden for T1D than the general population.

Our data also supports a role for genetic risk factors for T2D, especially in those who are obese where the relationship between genetic risk and diabetes may be modified by adiposity, with stronger effects observed among obese individuals (17). The significance of the T2D-PRS finding in obese individuals is that it demonstrates a genetic contribution above and beyond BMI alone, suggesting that genetic risk modifies the relationship between obesity and post-AP dysglycemia.

The dual elevation of both T1D-GRS and T2D-PRS challenges traditional diabetes classification and indicates that post-pancreatitis diabetes represents a heterogeneous condition. Some patients likely develop predominantly autoimmune diabetes (high T1D-GRS), while others may experience primarily metabolic dysfunction (high T2D-PRS), and many will show similar overlapping features. This heterogeneity has important therapeutic implications, as immunomodulatory approaches may benefit the autoimmune subset, whereas metabolic interventions may be more appropriate for others (18,19). Limitations of this study include its single-center design and small sample size. Furthermore, the use of UK Biobank as a population-level baseline introduces potential age-related biases as T1D is more prevalent in pediatric populations, the enrichment observed in our cohort may partially reflect this demographic difference rather than being specific to a history of pancreatitis. Additionally, survivor bias within the adult UK Biobank cohort might deflate its T1D-PRS proportions, as individuals with the highest genetic risk may be under-represented. Finally, whilst T1D-GRS comparisons followed standardized frameworks (13, 15), equivalent validated cutoffs for T2D-PRS were unavailable, necessitating caution in cross-study interpretations of these findings.

Future studies are needed to validate our findings by expanding recruitment and genotyping to improve statistical power, especially for subgroup analyses (e.g., AAb positivity and BMI categories). Ultimately, integrating genetic risk scores for T1D and T2D, especially for T1D, into clinical care for pancreatitis pediatric AP and expanding this knowledge to chronic pancreatitis patients may enable precision medicine approaches that reduce the diabetes burden in this vulnerable population.

## Abbreviations

AP: Acute Pancreatitis
GRS: Genetic Risk Scores
PRS: Polygenic Risk Score
T1D: Type 1 Diabetes
T2D: Type 2 Diabetes
UKBB: UK Biobank
GAP: Genetic Ancestry Prediction
MAF: Minor Allele Frequency
pPRS: partitioned PRS.

## ACKNOWLEDGEMENTS

The authors thank the patients and their families for their contributions to this study.

## ETHICAL APPROVAL

This study was approved by the Cincinnati Children’s Hospital Medical Center IRB panel IRB#2021-4050.

## DATA AVAILABILITY

All data generated or analyzed during this study are available upon reasonable request. All utilized SNP sets are open-source and directly downloadable through the PRSedm suite.

## AUTHOR CONTRIBUTIONS

A.L.G. and M.A.E.-H. were involved in the conceptualization of this study. L.Z., S.A.S., H.S., A.L.G., and M.A.E.-H. designed the study and contributed to the analysis and interpretation of the results. L.Z. and F.A. conducted the analyses. F.A., S.T., and C.A.W. collected and provided the data for this study. L.Z., F.A., A.L.G., and M.A.E.-H. wrote the first manuscript draft. All authors edited, reviewed, and approved the final version of the manuscript. A.L.G. and M.A.E.-H. are the guarantors of this work and, as such, had full access to all the data in the study and take responsibility for the integrity of the data and accuracy of the data analysis.

## SUPPLEMENTARY FIGURES

**Supplementary Figure 1.**
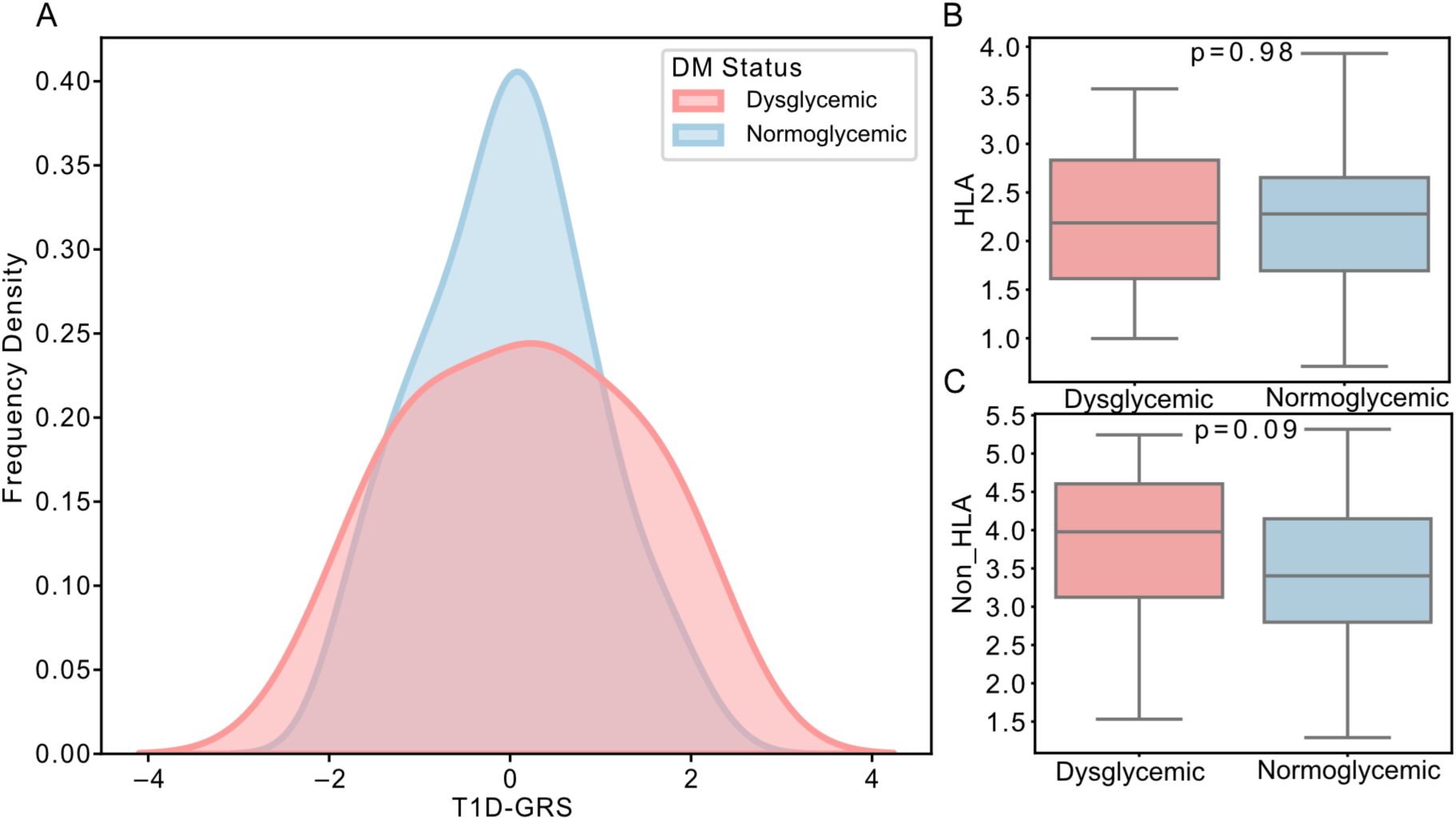
T1D-Genetic Risk Score (GRS) analysis by diabetes status. The analysis was conducted across a multi-ancestry population. **(A)** Frequency distribution of T1D-GRS. Participants are stratified into dysglycemic cases (prediabetes/diabetes; n=23) and normoglycemic controls (n=99). Differences between groups were analyzed using the Welch’s t-test (p = 0.57) **(B–C)** Partitioned GRS distribution for (B) HLA variants and (C) Non-HLA variants, compared by diabetes status. p-values displayed above each plot denote the statistical significance of differences.

**Supplementary Figure 2.**
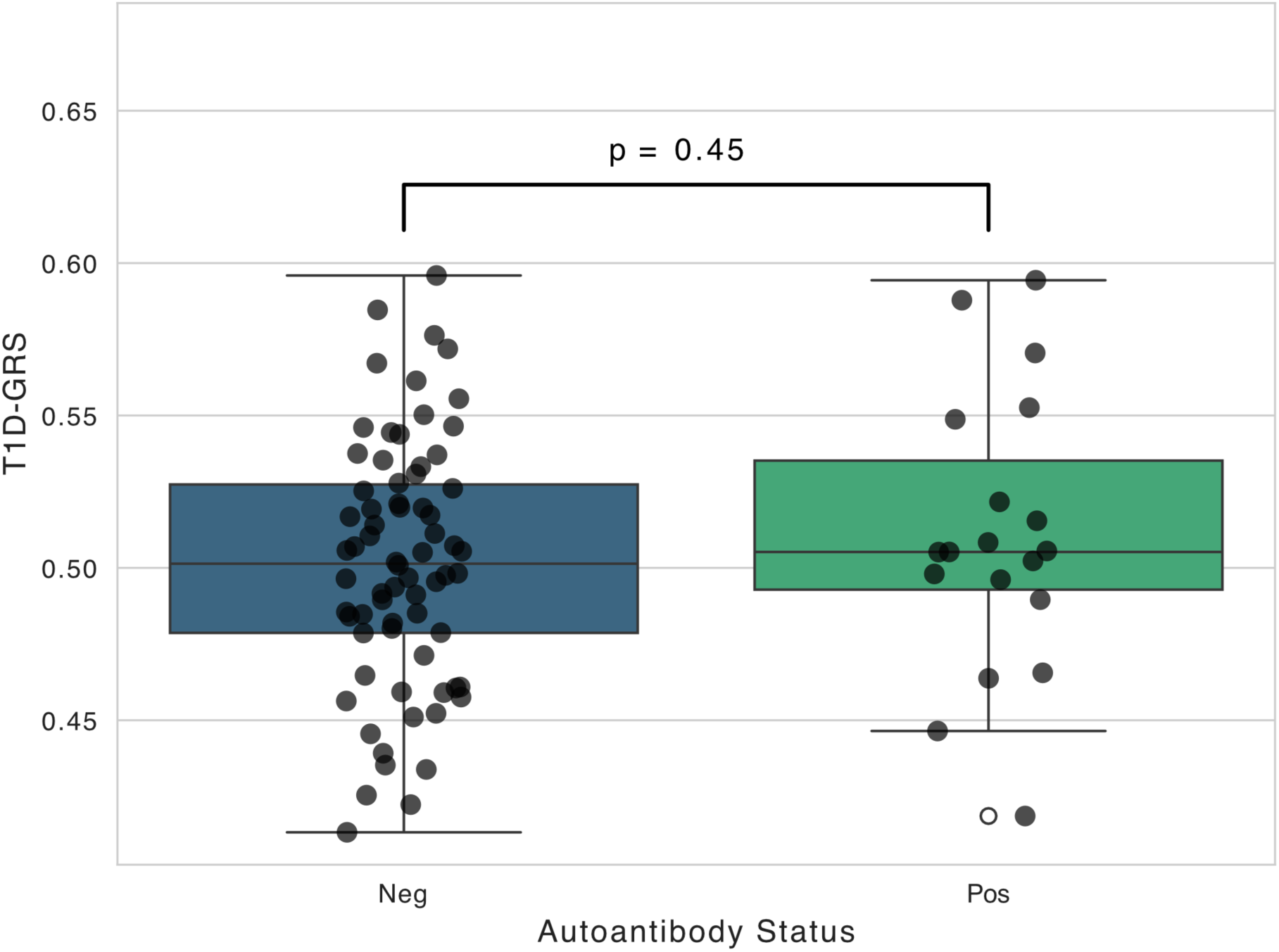
Comparison of T1D-Genetic Risk Score (GRS) by autoantibody status. Boxplots illustrate the distribution of T1D-GRS among autoantibody negative (Neg, n = 70) and positive (Pos, n = 19) participants in the full cohort. p-value indicated above the plot.

**Supplementary Figure 3.**
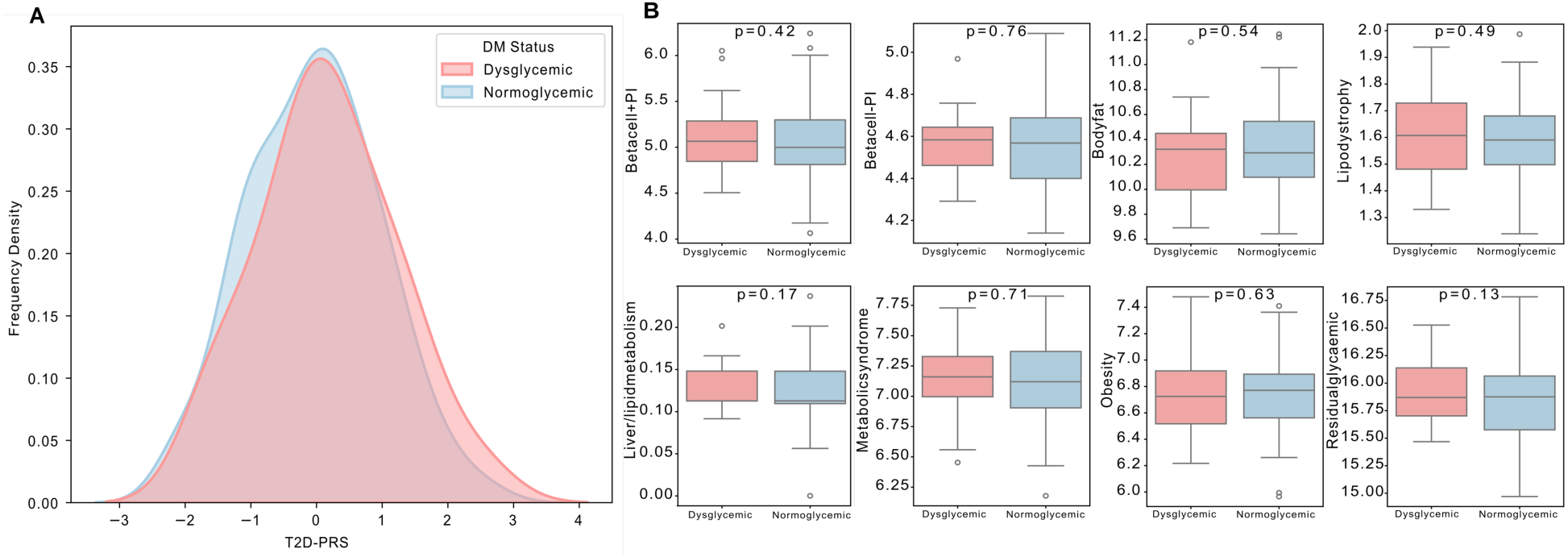
Comparison of T2D-PRS and partitioned components by diabetes status. The analysis was conducted across a multi-ancestry population. The global T2D-PRS was calculated using the t2dp-suzuki24-ma model from ancestry-specific hard clustering (14). Partitioned polygenic scores (pPS) were estimated through hard-clustering analyses to represent specific biological pathways. **(A)** Frequency distribution of T2D-PRS. Participants are stratified into dysglycemic cases (prediabetes/diabetes) and normoglycemic controls. Differences between groups were analyzed using the Welch’s t-test (p = 0.41) **(B)** Boxplots comparing partitioned PRS components between groups, with p-values indicated above each plot.

**Supplementary Figure 4.**
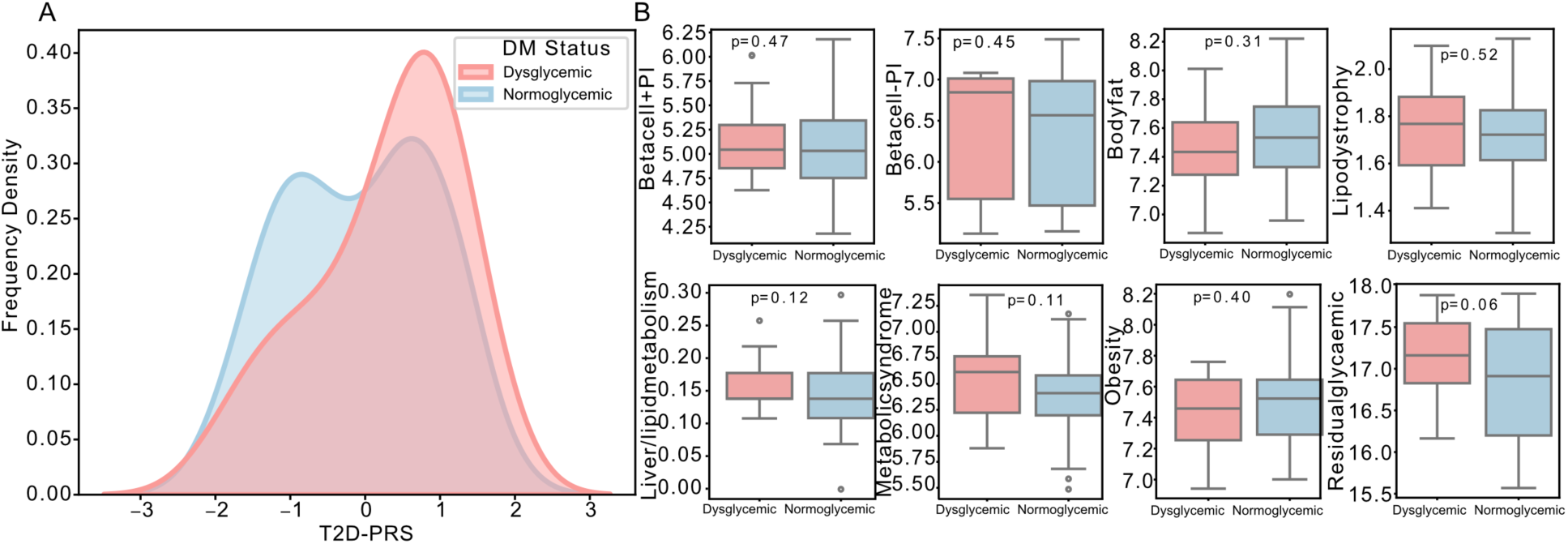
Comparison of T2D-PRS and partitioned components by diabetes status. Analysis restricted to the European (EUR) population. The global T2D-PRS was calculated using the t2dp-suzuki24-eur model from ancestry-specific hard clustering (14). Partitioned polygenic scores (pPS) were estimated through hard-clustering analyses to represent specific biological pathways. **(A)** Frequency distribution of T2D-PRS. Participants are stratified into dysglycemic cases (prediabetes/diabetes; n=20) and normoglycemic controls (n=72). Differences between groups were analyzed using the Welch’s t-test (p = 0.18). **(B)** Boxplots comparing partitioned PRS components between groups, with p-values indicated above each plot.

